# Variational autoencoders to predict DNA-methylation age and provide biological insights in age-related health and disease

**DOI:** 10.1101/2023.07.07.23292381

**Authors:** Sandra Steyaert, Adriaan Verhelle, Wim Van Criekinge

## Abstract

**Motivation:** Epigenetic ageing clocks based on age-associated changes in DNA-methylation (DNAm) profiles have shown great potential as predictors to quantify age-related health and disease. Most of these clocks are built using penalized linear regression models with the aim to predict chronological and/or biological age. However, the precise underlying biological mechanisms influencing these clocks remain unclear. In this work, we explored if a Variational Autoencoder (VAE) can be trained to model DNAm age, and whether this VAE captures biologically relevant features of ageing in the latent space.

**Results:** Our results indicate that VAEs present a promising framework to construct embeddings that capture complex interactions and that can be used to extract biological meaningful features upon predicting DNAm age. By using deep learning interpretation methods, we show it is possible to determine which genomic loci and pathways are important in making the predictions, both on a population and individual level, paving the way to unravel what makes the DNAm clock tick.

## Introduction

Ageing is a gradual biological process characterized by functional decline in both physical and cognitive capabilities. It represents a major risk factor for age-related disorders such as neurodegenerative disease, rheumatoid arthritis, osteoarthritis, cancer, diabetes and cardiovascular diseases, and ultimately increased risk of death^1^. The past two decades, a growing interest in longevity (lifespan) and healthy ageing (healthspan) led to tremendous efforts to unravel the biological basis with the hope to delay or prevent age-related conditions and increase both life-and healthspan^2^. Ageing is regulated by complex cellular and molecular mechanisms, and variation in life-and healthspan between humans is affected by a variety of factors including genetic background but also socioeconomic and environmental elements^3^.

Epigenetics connects the genotype to the phenotype and plays an important part in the response to environmental factors hinting at a lead role in modulating the ageing process^4, 5^. Multiple studies indeed have shown the link between rate of ageing and alterations in epigenetic regulators such as DNA-methylation (DNAm), histone modifications, non-coding RNAs and chromatin organization^6, 7^. As epigenetic modifications are potentially reversible, these findings greatly facilitated the emerging anti-ageing field and the development of ageing-delaying or even reverse-ageing therapies^8, 9^. Especially DNAm levels have been the focus of many age-related studies as DNAm has been well-reported to dynamically change with age^5^. For example, DNAm levels tend to increase with age at some CpG islands whereas loci outside CpG islands usually lose methylation^6^.

Interestingly, these studies further showed that the DNA methylome, and in particular specific collections of individual 5-methylcytosine sites, can be used to predict chronological age of a variety of tissues^10, 11^, and in 2013 Horvath introduced the concept of the “epigenetic or DNAm clock”^12^. Furthermore, apart from being a predictor for chronological age, DNAm may also serve as a valuable biomarker for healthy versus unhealthy ageing and disease risk, and thus a proxy for “biological age”^13^. Importantly, while chronological age is independent of genetic background, lifestyle, health and disease, biological age can be highly variable between people of the same chronological age. Indeed, disparity between an individuals’ chronological and biological age may reflect the impact of specific genetic and environmental factors, leading to an acceleration or deceleration of age-related functional decline compared to peers of the same chronological age^14, 15^. Multiple methods have been proposed to measure biological age, including physiological^16^ (e.g. blood, cardiovascular, cognitive and physical parameters) as well as some molecular (e.g. telomere length^17^, mitochondrial function^18^) markers. However, despite its potential as an objective quantification of healthspan, there is currently no consensus on the best method to determine biological age^19, 20^, mainly reflecting its inherent complexity. However, some recent studies have stipulated that the delta between one’s DNAm age and true chronological age, might be a surrogate for biological age^13, 21^. In this regard, a positive delta indicates an DNAm age-acceleration resulting in a biologically older individual, while a negative delta reflects that the individual aged slower and is biologically younger. For example, one study investigating blood of centenarians and their offspring reported a consistent negative delta, i.e. DNAm age-deceleration, for this population^22^.

In light of these findings, there has been broad interest in the use of DNAm age. This, together with the advances in DNAm technologies and the data availability, enabled the creation of several methylation clocks^23^. The first methylation clocks were created for humans but have recently also been developed for other organisms like mice and dogs, amongst others. The best-known human DNAm clocks are probably those from Horvath^12^ (multi-tissue), Hannum^24^ (blood), Levine^13^ (PhenoAge, blood) and Lu^25^ (GrimAge, blood) but many others exist. Here, the former two are referred to as first-generation clocks because they were trained to predict chronological age. The latter two clocks, on the other hand, are labelled second-generation clocks, because they were trained on a surrogate biomarker reflecting ageing phenotypes to better reflect disease morbidity and mortality with the aim to predict biological age.

While each clock slightly differs in the used datasets and/or data processing approach, the vast majority are linear models of a set of CpGs, with or without intercept, regressing to either chronological age or to a proxy for biological age. Each linear model is typically built using the same general supervised workflow, namely by leveraging a penalized or regularized regression model like ElasticNet or Lasso to automatically select the most informative set of CpGs for the prediction out of the full methylation array after which a linear model is developed on those CpGs^14, 26^. The final number of selected CpGs differs for each linear model but usually ranges between 3 and 2000. For example, the Horvath, Hannum, Levine and Lu clocks contain 353, 71, 513 and 1,113 CpGs respectively^27^. While regularized linear regression has been widely used to develop epigenetic clocks, there are some important limitations with this approach. Not only can linear regression display high bias, but it is also not able to capture more complex, non-linear CpG interactions^28^. More importantly, epigenetic clocks based on linear models assume that the contribution of each CpG is additive and that the rate of methylation change is constant over the entire lifespan. In addition, regularized regression methods are often sensitive to subtle differences in the used data. This is evidenced by the fact that many of the published clocks use completely different, non-overlapping sets of CpGs, even when trained for the same tissue, making it difficult to assess any biological or functional connection to the ageing process^26^.

Deep learning (DL) has proved to be a powerful and flexible approach for many data types, including molecular data^29–32^. When provided with large, high-dimensional data, neural networks (NN) can capture complex linear and non-linear feature interactions. Also, for DNAm data and DNAm age prediction, NN models have already been successfully applied. For example, methods like DeepMAge^33^ and AltumAge^28^ use a deep NN for blood and pan-tissue DNAm age prediction, respectively. By using a multi-layer perceptron (MLP) architecture, both methods outperform regularized linear regression models such as Horvath in chronological age prediction, seem to be more resilient to noise and generalize better to new data. Furthermore, they illustrate that NNs allow to simultaneously use information of thousands of CpGs, can capture higher-order, complex feature interactions and that it is possible to determine the contribution of each CpGs by leveraging DL interpretation methods like Shapley Additive Explanation (SHAP)^34^. As such, these results show that DL is a very encouraging approach for epigenetic clock predictions and to unravel complex biological mechanisms within the context of ageing^35^.

Among DL models, variational autoencoders (VAE) are an emerging technique that hold great potential in a wide range of applications. VAEs are data driven, unsupervised generative models that harness the power of DL and learn data distributions without the need for accurate labels^36^. The underlying architecture consists of autoencoding layers which first encode (compress) the data in a lower-dimensional latent space after which the data is decoded (reconstructed) into its original dimension. Here, the aim of the encoder phase is not only to reduce the data dimensionality, but to compress the data by removing redundant information while at the same time keeping the most important information relevant for the research question in this reduced representation^37^. While a traditional AE minimizes the reconstruction error during training and results in a non-regularized latent space (the decoder cannot be used to generate valid input data from vectors sampled from the latent space), a VAE is stochastic and learns the parameters of the data distribution, i.e. mean and standard deviation, ensuring a latent space with generative capabilities^38^. Importantly, the fact that the autoencoding makes use of complex, non-linear activation functions, allows the VAE to learn complex, lower-dimensional representations of the data. Because of the ability to apprehend underlying data manifolds, VAEs have been employed to generate meaningful biological latent spaces for various biological applications. For instance, Way *et al.* trained a VAE on TCGA pan-cancer RNA-seq data to model cancer gene expression and observed that the encoded lower-dimensional features contained biological meaningful signals^39^. Similarly, Kinalis *et al.* show that it is possible to directly deconvolute biological modules inherent in single cell RNA-seq data that outline cell-specific drivers, without making any prior assumptions^40^. Of late, VAEs are being used to reduce the dimensionality of DNAm data.

Methylation profiles are typically measured via DNAm microarrays such as Illumina’s HumanMethylation27 (27K), HumanMethylation450 (450K) and the most recent HumanMethylationEPIC (850K), measuring approximately 27,000, 450,000 and 850,000 CpG sites, respectively, and are reported as continuous beta-values ranging from 0 (no methylation), to 1 (fully methylated)^41^. Given the high number of features, VAEs have been applied to embed this data to into a lower-dimensional meaningful feature set^42–44^. For example, Levy *et al.* investigated the use of VAEs on six different DNAm datasets and demonstrated its potential to capture cancer subtype information, cellular differences, and smoking factors, and to make age predictions^44^. Two other DNAm breast cancer studies illustrated the use of the encoded latent space to predict cancer subtypes^42, 43^. In addition, when comparing to regularized regression-based methods for feature selection, VAEs were able to reveal novel information about DNAm patterns in breast cancer^42^.

Here, we explore the use of VAEs to create a biologically relevant latent space that can be used to extract relevant biological signals related to ageing but also to model DNAm clocks to predict biological age. For this, we developed framework that first performs a prior dimensionality reduction to ∼13k CpGs which are fed into a VAE model. In a next step, the learned latent space is used as input of an MLP to predict age. We trained our framework using 8 publicly available DNAm 450K datasets totaling 2,716 blood samples originating from healthy individuals with chronological ages ranging between 0-103. We present initial results from using VAEs as a dimensionality reduction method for DNAm ageing applications and show that this lower dimensional latent space holds relevant information that may lead to mechanistic hypotheses on how the methylome drives the ageing process. We further demonstrate that this learned latent space can subsequently be used as input features for models to estimate age. Additional validation on two independent cohort not only illustrates that the developed models can be used to calculate DNAm age, but that the VAE has the potential to be used to distinguish diseased from health samples for age-related disorders.

Taken together, our analysis hints towards more robust DL-based methylation clocks with better estimates for biological age that at the same time can unravel complex biological networks involved in ageing and/or age-related disorders. It is our expectation that in the future, VAEs or other DL feature selection methods that can capture complex biological functions pave the way to combine multiple modalities relevant for ageing, e,g. molecular (DNAm, gene expression, metabolic profiles – single or multi-tissue), image data as well as socioeconomic factors and health records in one multimodal model for biological age prediction.

## Methods

### Data & Preprocessing

Methylation data used in this study consisted of 2,716 blood samples measured on Illumina’s Infinium HumanMethylation450 BeadChip (450K) array originating from seven public datasets. Raw data was downloaded from the Gene Expression Omnibus (GEO)^45^ (six datasets, i.e. GSE30870^46^, GSE32149^47^, GSE41169^48^, GSE36064^49^, GSE40279^24^, GSE87571^50^) and ArrayExpress^51^ (SATSA dataset^52^ - E-MTAB-7309). Details on the individual datasets and characteristics of the specific study cohort can be found in **Table 1**. A full description of each dataset can be found in the original reference.

**Table 1:** Overview of used 450K methylation datasets.

| Dataset | #Samples (origin) | Description | Median Age (min - max) |
| --- | --- | --- | --- |
| GSE30870<sup>46</sup> | 40 (blood PBMC) | The study aimed to compare the DNAm differences between newborns and nonagenarians using methylation array technology | 44.5 (0-103) |
| GSE32149<sup>47</sup> | 46 (blood PBMC) | DNAm study of Crohn's disease (n=14), ulcerative colitis (n=8) and controls (n=14). DNAm age was not found to be associated with disease status. | 15 (3.5-76) |
| GSE41169<sup>48</sup> | 95 (whole blood) | Genome wide DNAm profiling of whole blood in schizophrenia patients and healthy subjects of different ages. Dataset included 62 schizophrenia patients and 33 healthy subjects from Dutch descent. | 29 (18-65) |
| GSE36064<sup>49</sup> | 78 (blood PBMC) | DNAm association study in healthy children (males) | 3.1 (1-16) |
| GSE40279<sup>24</sup> | 656 (whole blood) | Genome wide DNAm profiling of healthy individuals across a large age range. | 65 (19-101) |
| GSE87571<sup>50</sup> | 729 (whole blood) | Genome wide DNAm profiling of healthy individuals across a large age range. | 47 (14-94) |
| SATSA<sup>52</sup> | 1072 (blood PBMC) | DNAm of longitudinal samples from The Swedish Adoption/Twin Study of Aging (SATSA). ~400 Swedish twins were sampled over 20 years (up to 5 times per sample). | 73.5 (48-99) |

The Illumina 450K array measures bisulfite-conversion-based, single-CpG resolution DNA methylation levels for over 480K CpG sites and covers 96% of CpG islands in the human genome^53^. Unlike the previous 27K platform, the Illumina 450K array includes two distinct probe types, i.e. Infinium I (n=135,501) and Infinium II (n=350,076). In the Infinium type, each CpG cute is targeted by two 50bp probes: one for detecting the methylated intensity (M) and one for detecting the unmethylated intensity (U). In contrast, Infinium II probes detect both the M and U intensity of each CpG site by one single probe using different dye colors (green and red). For each CpG site, methylation values are indicated by the beta-value which ranges from 0 (no methylation) to 1 (fully methylated) and is calculated as beta=M/(M+U+alpha) where alpha generally equals 100^41, 53^.

Raw beta-values were processed in R (v4.1.2) with the RnBeads package (v2.12.2). Probes not in CpG context were filtered out as well as probes for which the beta-values were NA or had low variability (standard deviation < 0.005). Beta values of the remaining probes were next normalized using the Beta Mixture Quantile dilation method (BMIQ) with an Exponential-Normal mixture (Enmix) signal intensity background correction. After preprocessing and normalization, only probes present in all eight datasets were kept resulting in a final dataset of 2,716 samples and 415,594 CpG probes. **Table 2** summarizes the number of samples per age group for this population.

**Table 2:**
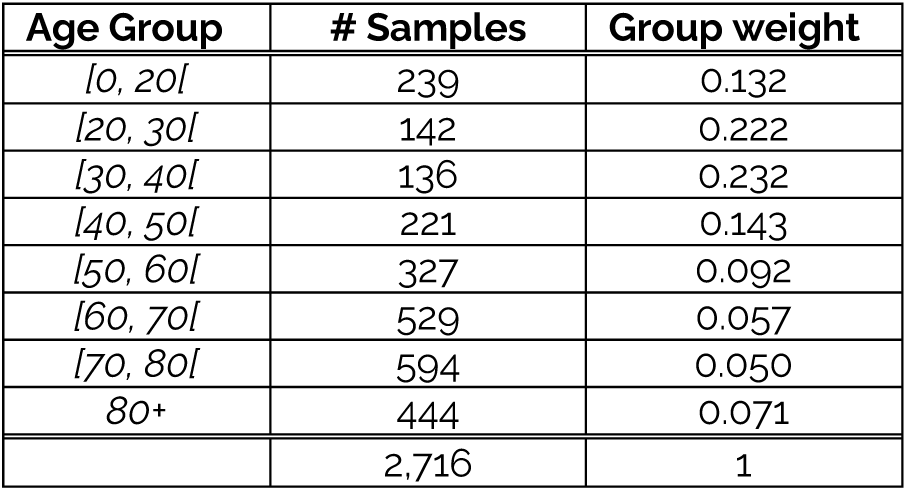
Number of samples per age group with their respective weights.

### Dimensionality Reduction

Typically, methylation arrays inherently present a dimensionality imbalance (i.e. number of CpGs (p) >> number of samples (n)). To handle the dimensionality imbalance in our dataset (p = 415,594, n = 2,716), a prior feature selection step was performed using ElasticNets, a shrinkage technique that uses a weighted combination of L1 (Ridge) and L2 (Lasso) regularization. As shown in **Table 2**, the predictor variable age is not uniformly distributed our dataset, i.e. the samples are imbalanced between different age classes. Therefore, before fitting an ElasticNet, samples were bootstrapped with a fraction of 42% and sample weights equal to the respective age class weight (**Table 2**). **Figure 1** shows the original age density distribution (panel **A**) and the resulting density distribution of one such bootstrap (panel **B**).

**Figure 1:**
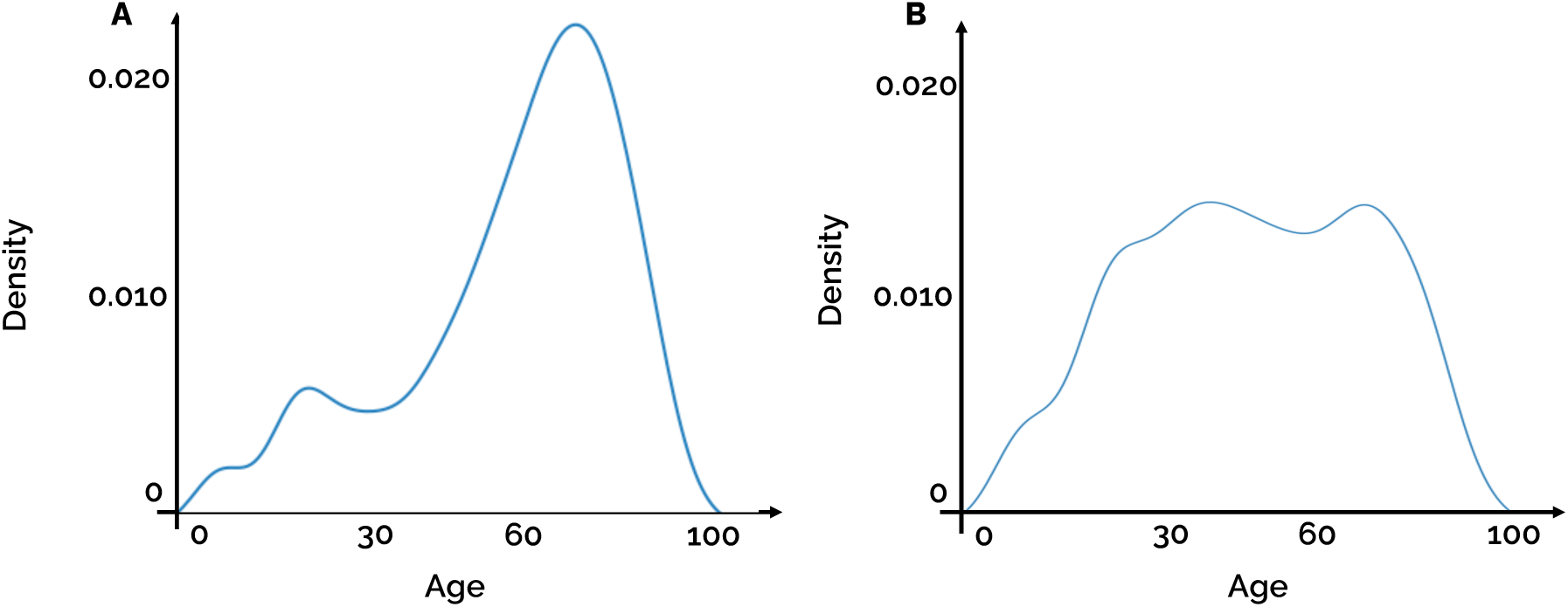
Age distribution of sample population. (A) original age density distribution. (B) age density distribution of first weighted bootstrap.

This bootstrapping procedure was performed 42 times, and for each bootstrapped distribution an ElasticNet was fitted with an L1/L2 ratio of 0.5 and the non-zero coefficients stored. On average, each ElasticNet had 895 non-zero coefficients with a range of [821; 955]. In a next step, the non-zero coefficients from all bootstrap iterations were combined, resulting in 13,242 unique non-zero coefficients, i.e. CpGs, out of the initial 415,594 (=3.19%).

### Model Development

#### Variational Autoencoder Model

We employed a variational autoencoder (VAE), an unsupervised deep learning architecture that consists of two neural network parts: an encoder and a decoder (**Figure 2A**). The encoder converts the input (our MxN data matrix with M=2,716 samples, and N=13,242 CpGs) to a lower dimensional latent space consisting of two feature vectors representing the input as a jointly Gaussian distribution. The decoder then randomly samples from this latent distribution to regenerate the data. Here, the effectiveness of the model is evaluated by comparing reconstructed data to the original input data. Importantly, the latent space (also called embedding) is the key concept of the VAE: each of the input features is combined in a weighted (nonlinear) pattern that contributes to the learned latent representation^36^. Hence the encoder can be seen as a complex feature extractor selecting relevant signals in the data while removing the noise.

**Figure 2:**
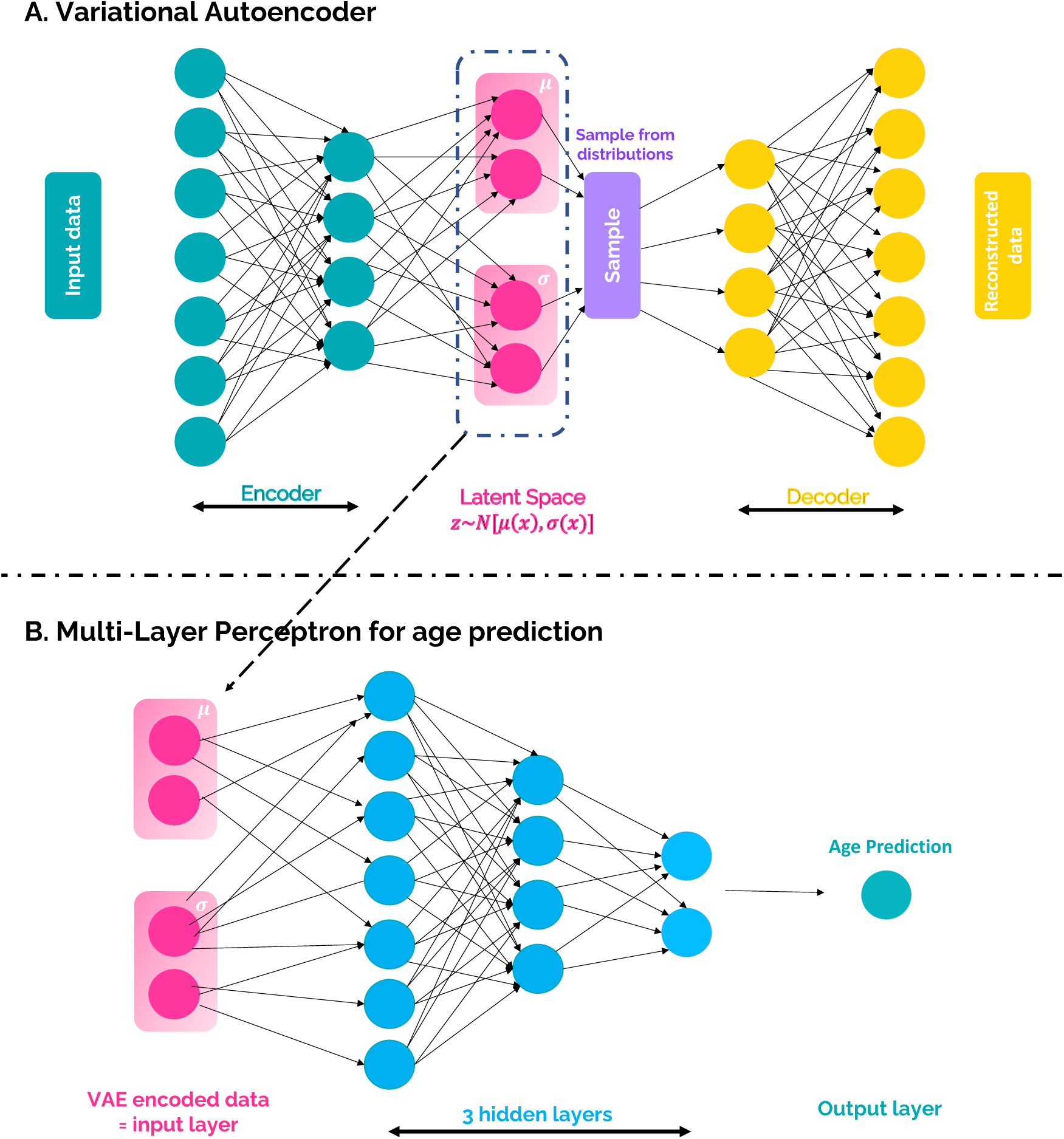
Overview of Model Architecture. (A) Variational Autoencoder (VAE) to embed the methylation data into a latent space. (B) Three-layer Multi-Layer Perception (MLP) for age prediction using the VAE embedding as input features.

We constructed a VAE with 2 linear layers for encoding with dimensions 4096 and 2048 respectively, a latent space of size 242 and a two linear decoding layers with dimensions 2048 and 4096, respectively (**Figure 2A)**. Between each linear layer, a 1D batch normalization was performed and the non-linear ReLu function was used as activation function. The input of the VAE was limited to the 13,242 CpGs selected by the ElasticNet analysis. The VAE model was trained using the ADAM optimizer^54^ with two loss functions: (i) a Mean Squared Error (MSE) loss function capturing the reconstruction loss and (ii) a Kullback-Leibler divergence (KLD) loss function measuring the distance between two distributions. Two KLD implementations were tested: (i) one that determines the KLD between the latent distribution and a Gaussian distribution, thereby assuming hence forcing the sampling in the representation layer to approximate a normal random variable, and (ii) a Monte Carlo approximation of KLD which is distribution-agnostic, i.e. without making any prior assumptions about the underlying distribution^55^. The total training loss was defined as: MSEloss + *w*\*KLDloss with *w* the weight of the KLD term varying between 0-1 following a cyclical sigmoid annealing scheduling to mitigate KLD-vanishing^56^.

Samples were shuffled in a training and test set at an 80/20 ratio with stratification on age class. This test set was left out during model training and only used for final model performance. A 10-fold cross validation (CV) performed on the training set. Hyperparameters were empirically determined. Each CV model was trained for 100 epochs using a batch size of 64, a learning rate of 1e-3. To prevent exploding gradients during VAE training, a weight decay (L2 regularization) of 1e-4 was set and gradients were clipped with a norm of 0.01. For each CV, the weights were saved for the epoch with the lowest loss on the validation set. The best model was chosen based on the CV configuration with the highest validation accuracy. This final model was next evaluated on the test set.

#### Multi-Layer Perceptron

In order to test the utility of the learned latent methylation space to predict age, a three-layer multi-layer perceptron (MLP) was trained with input layer of size 242 (= size of latent space), 3 hidden layers of size 128, 64 and 32 and output layer of size 1 (**Figure 2B**). Before each layer, a dropout with probability 0.42 was implemented and the non-linear ReLu function was used as activation function. As for the VAE, the ADAM optimizer was used together with a weighted MSE loss function, where the error on each sample was weighted according to its age class weight (**Table 2**). The MLP was trained for 100 epochs, using a batch size of 32, learning rate of 1e-3 and weight decay of 1e-4. Also here, the best model was selected as the epoch with the highest performance on the validation set and evaluated on the test set.

### Model Validation

To evaluate the VAE and MLP models, additional validation was done on two independent datasets. A first dataset consisted of 418 DNAm samples originating from an African American sibling cohort from the Genetic Epidemiology Network of Arteriopathy (GENOA) study (GEO accession GSE210254)^57^. DNAm age was calculated for each sample, and performance evaluated by calculating the mean absolute error (MAE) and r^2^. Results were further compared to Horvath, Hannum and PhenoAge DNAm age predictions.

A second dataset consisted of 689 peripheral blood leukocytes samples from a DNAm study to determine whether rheumatoid arthritis (RA) patients have methylation differences comparing to normal controls (GEO accession GSE42861)^58^. Here, although with a small effect size (age difference of 1.2 years), the researchers found a significant (p=0.0049) difference between the two groups^58^. While Horvath did not find a significant difference of negative age acceleration between the diseased versus healthy samples^12^, we wanted to evaluate if our models could pick up any clinical signal or deviation in RA patients. To test this, we first computed a universal measure of age acceleration (AgeAcc) as the DNAm age predicted by our model divided by chronological age. We also calculated AgeAcc using DNAm age predicted by Horvath, Hannum and PhenoAge clocks.

To assess if the latent space encoded by our VAE can be used as a biological feature extractor for age-related features, we trained a simple logistic regression model to predict RA status (**Figure 3**). The dataset was split in an 80/20 ratio with stratification on RA status and a Logistic Regression (Logit) model was trained on the training set using sklearn’s default LogisticRegressionCV function with L1 penalty and the saga solver. Performance was measured on the test set by calculating the confusion matrix and generating the Receiver Operating Characteristic (ROC) curves.

**Figure 3:**
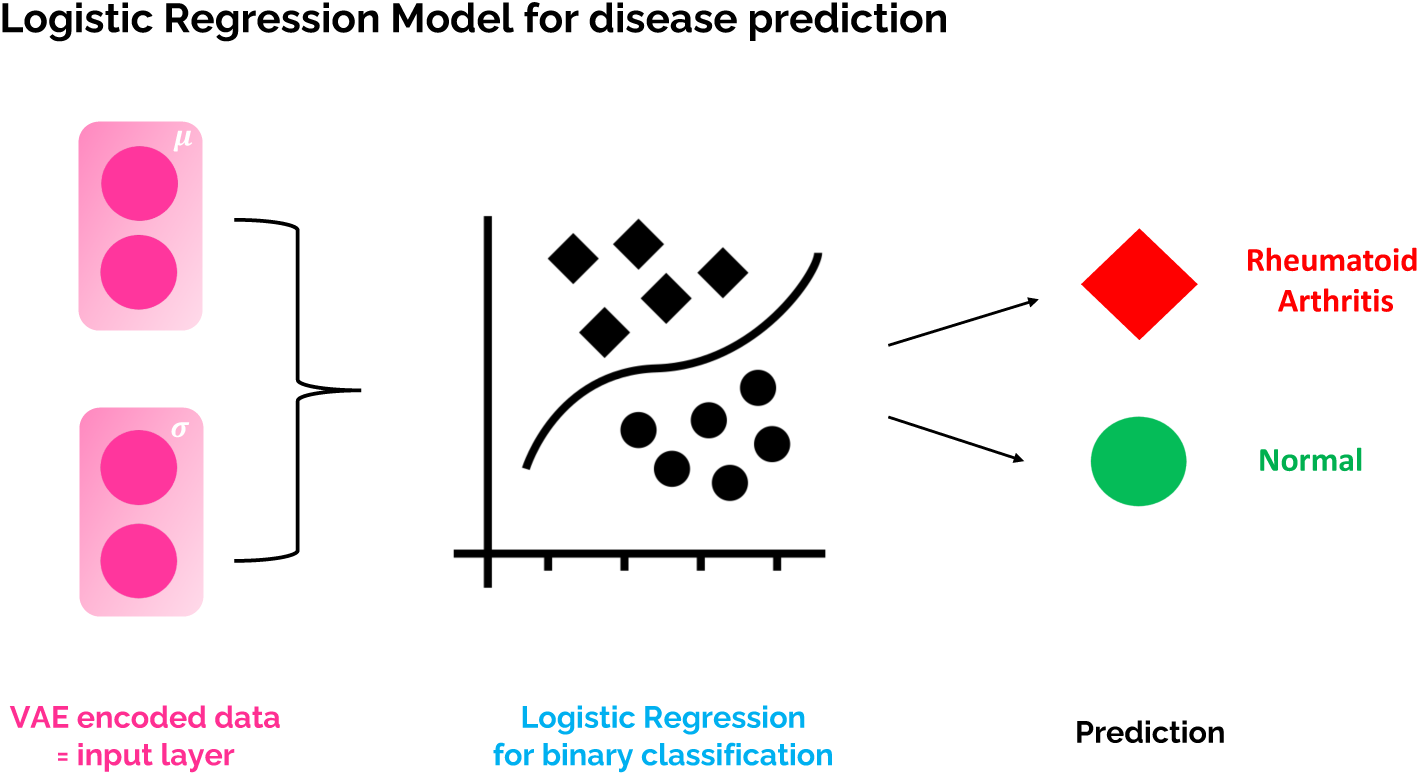
Logistic regression model for binary classification to predict RA status from the VAE embedding.

These datasets were not used in any training or fine-tuning step but kept as a separate independent cohort for final model validation.

### Model Interpretation

While DL can extract predictive features from complex data, these are usually abstract. Here, we propose a method that leverages the VAE to biologically interpret the latent space in a post-hoc manner. The relation with the input features (13,242CpGs) was determined by backpropagation of the VAE model and calculating the model gradients. The importance of each gene was next assessed by averaging the gradients of the gene-associated CpGs. Likewise, the importance of each pathway in each neuron was assessed by averaging the gene importances of the associated gene set. Here, gene sets were defined by mapping individual genes to the Reactome pathway collection (C2:CP collection v7.5, downloaded from https://data.broadinstitute.org/gsea-msigdb/msigdb/release/7.5/)^59, 60^.

A cluster analysis was performed by visualizing the latent space embeddings on a 2D and 3D space by calculating t–Stochastic Neighborhood Embedding (t-SNE) and Uniform Manifold Approximation and Projection (UMAP) projections using their respective python packages^61, 62^.

## Results

### Model development for epigenetic age prediction

Two VAE frameworks were developed and evaluated for epigenetic age prediction. The first framework employs a VAE with the KLD calculated between the latent distribution and a Gaussian distribution, while the second framework utilizes a distribution-agnostic approach for KLD calculation. For each framework, an MLP was trained using 10-fold CV to predict age using the learned latent spaces of the samples as input (**Figure 2**). The performance of the models was evaluated using two standard evaluation metrics: Mean Absolute Error (MAE) and the coefficient of determination, denoted as r-squared (r^2^). **Table 3** presents the CV model performances, including the mean score of the 10-fold CV with standard deviation for both frameworks on the training, validation, and test sets. Remarkably, the agnostic framework consistently outperformed the Gaussian framework in all datasets, exhibiting lower MAE and higher r^2^ values. Further statistical analysis, specifically a pairwise comparison using the Wilcoxon signed-rank test, confirmed the significantly improved performance of the agnostic framework over the Gaussian framework, as depicted in **Figure 4**.

**Figure 4:**
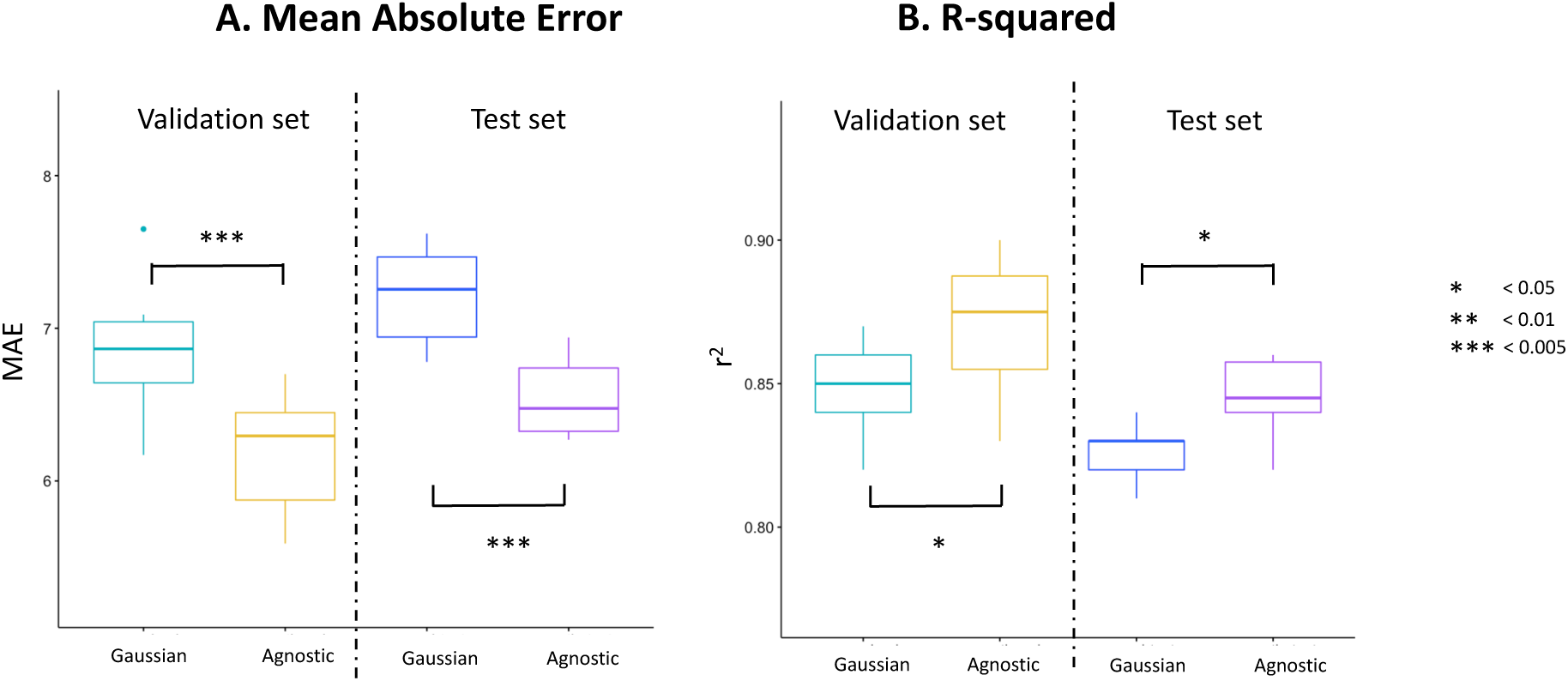
Boxplots illustrating the model performance of each Kullback-Leibler Divergence (KLD) strategy on the validation (N=218) and test (N=543) datasets. Subfigure (A) presents the distribution of Mean Absolute Error (MAE) obtained from 10-fold cross-validation (CV), while subfigure (B) displays the coefficient of determination (r2) scores for each CV fold. Statistical significance between the KLD strategies was assessed using pairwise Wilcoxon signed-rank test, with significance levels denoted by asterisks (*P value <0.05, **P value <0.01, and ***P value <0.005), indicating the level of significance in relation to the comparison of the two strategies.

**Table 3:** Performance analysis of the variational autoencoder (VAE) combined with a multilayer perceptron (MLP) framework for the prediction of epigenetic age. The table includes results of the Mean Absolute Error (MAE) and the coefficient of determination (r2) obtained from cross-validation (CV) on the training, validation, and test datasets. The performances are reported in terms of the mean values, along with the corresponding standard deviations (stdev), for both the Gaussian Kullback-Leibler Divergence (KLD) strategy and the distribution-agnostic KLD strategy.

| Dataset | Metric | KLD strategy |  |
| --- | --- | --- | --- |
|  |  | Gaussian | Agnostic |
| Training | Mean (MAE) CV<br>± stdev | 7.00<br>± 0.23 | 6.35<br>± 0.25 |
| | Mean($r^2$) CV<br>± stdev | 0.84<br>± 0.01 | 0.87<br>± 0.01 |
| Validation | Mean (MAE) CV<br>± stdev | 6.83<br>± 0.43 | 6.20<br>± 0.38 |
| | Mean($r^2$) CV<br>± stdev | 0.85<br>± 0.01 | 0.87<br>± 0.02 |
| Test | Mean($r^2$) CV<br>± stdev | 7.22<br>± 0.31 | 6.54<br>± 0.28 |
| | Mean($r^2$) CV<br>± stdev | 0.83<br>± 0.01 | 0.85<br>± 0.01 |

**Figure 5** showcases the predictions and evaluation metrics of the best-performing CV distribution-agnostic model for all samples included in the training, validation, and test sets. The x-axis represents the chronological age of each sample, while the y-axis displays the age predicted by the model. This final optimized model, referred to as methVAgE in subsequent analyses, achieved an MAE of 6.34 and an r^2^ of 0.86 on the test set.

### Model evaluation on external datasets

After pre-processing the methylation data of the GENOA^57^ (African American sibling cohort from the Genetic Epidemiology Network of Arteriopathy) and rheumatoid arthritis^58^ (RA) datasets, the developed model was subsequently validated on these two external cohorts.

**Figure 6** illustrates the predictions on 418 GENOA samples from three epigenetic clocks, namely (i) our VAE+MLP framework (referred to as methVAgE), and two other widely used linear model clocks, (ii) Horvath^12^ and (iii) PhenoAge^13^. Additionally, we calculated the age acceleration (AgeAcc) of each sample by dividing the predicted DNAm age by the chronological age. An AgeAcc value greater than 1 indicates accelerated biological ageing, whereas a value lower than 1 implies decelerated biological ageing, which is preferable. **Figure 6** also displays the mean AgeAcc of all samples for each clock.

**Figure 5:**
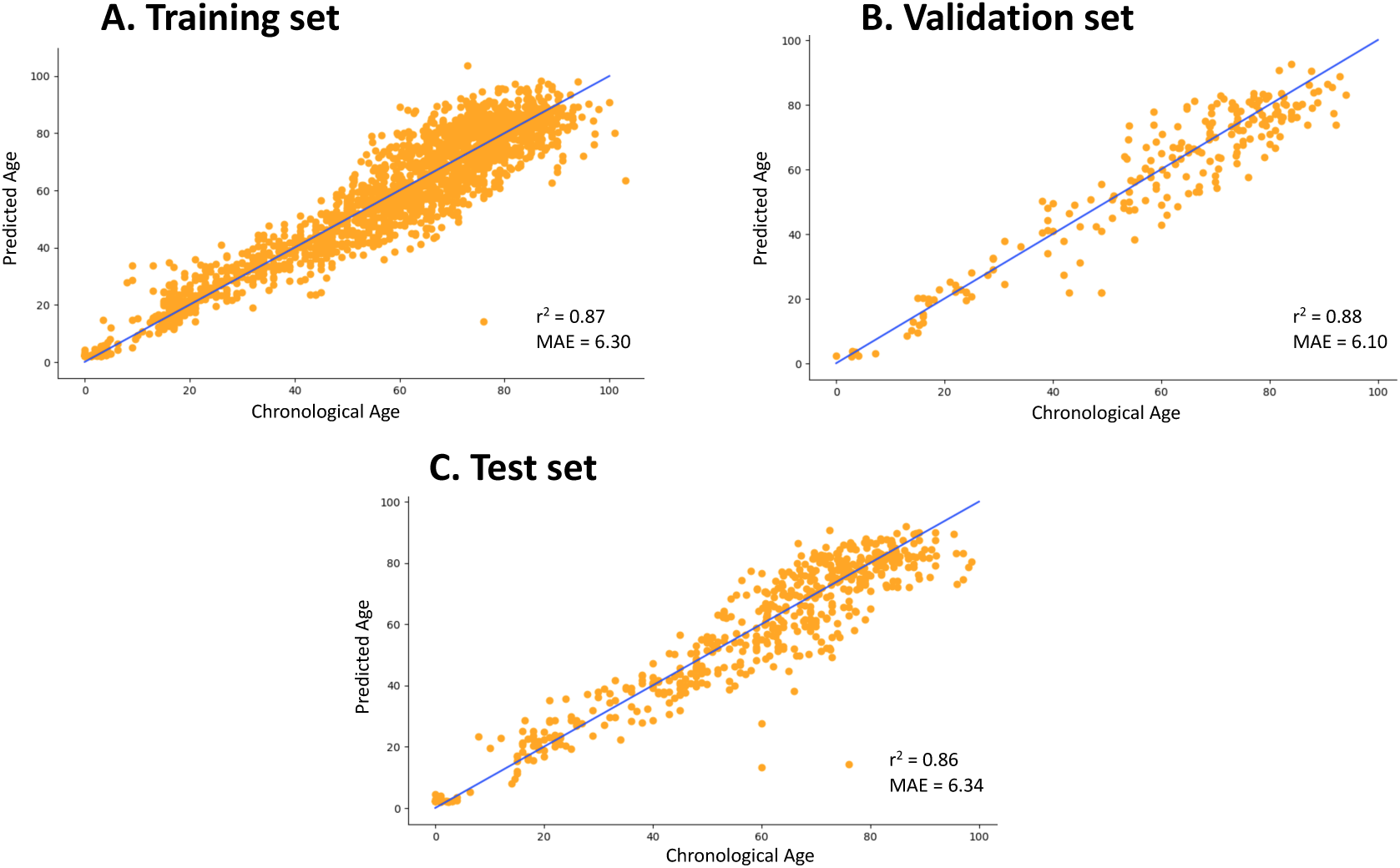
Predictions and evaluation metrics of the best-performing CV distribution-agnostic model for all samples in training (A), validation (B), and test (C) sets. For each panel, the x-axis represents chronological age, and the y-axis model-predicted DNAm age.

**Figure 6:**
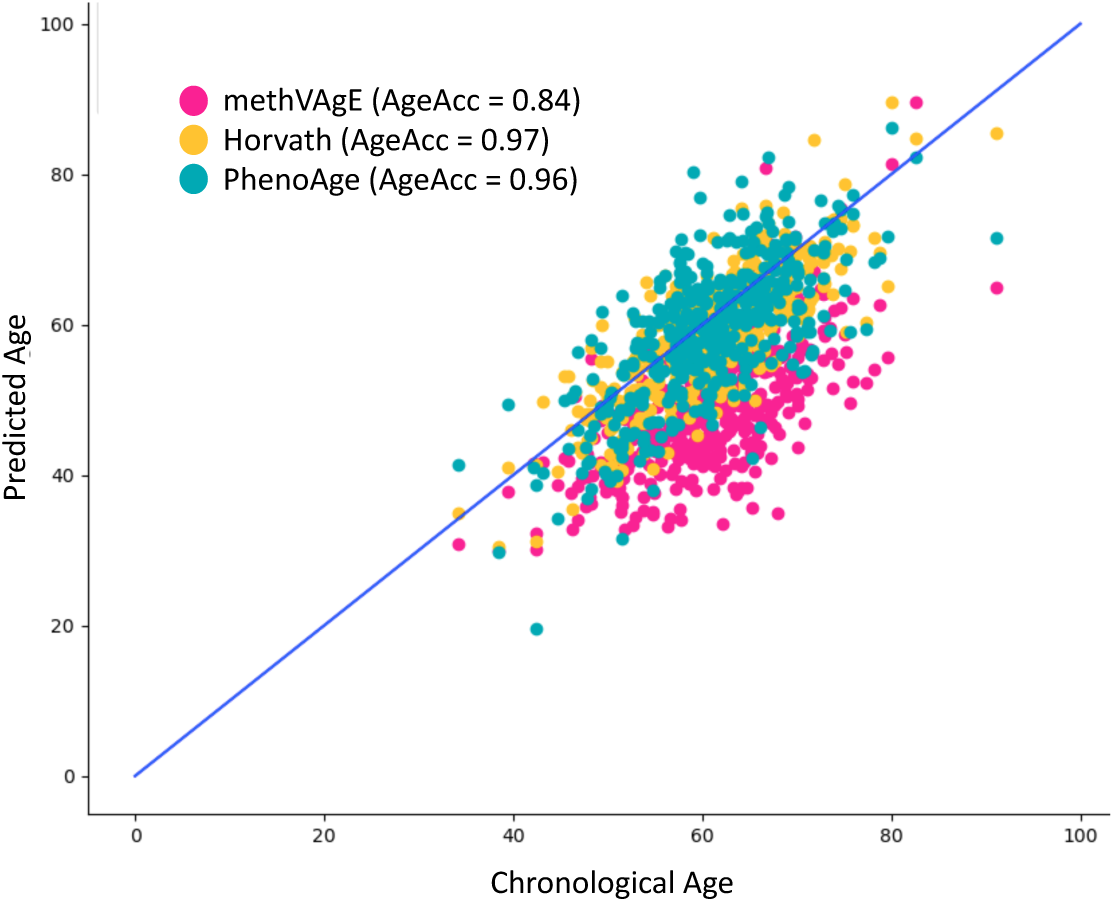
Epigenetic clock predictions on GENOA samples. Comparison of methVAgE (our VAE+MLP framework) with Horvath and PhenoAge (two widely used linear model clocks). X-axis represents chronological age, and y-axis the predicted DNAm age. The mean age acceleration (AgeAcc) - calculated as predicted DNAm age divided by chronological age averaged over all samples - is also displayed for each clock.

Notably, the DNAm age predictions of our methVAgE model consistently trend lower than the corresponding chronological ages at the cohort level, as evidenced by the lower AgeAcc compared to the other two clocks. This observation is intriguing considering previous findings^63^ demonstrating that African Americans tend to exhibit significantly lower extrinsic DNAm ages compared to whites. Although both Horvath and PhenoAge clocks also exhibit an average AgeAcc below 1 for this cohort, the effect is less pronounced, suggesting that our framework may better capture underlying biological signals. Additionally, we calculated the AgeAcc for Hannum^24^, another established epigenetic clock (not shown in **Figure 6**), which yields an average AgeAcc of 1.06, indicating that this biological phenomenon is not adequately captured by this particular clock.

**Figure 7** depicts the model predictions of the epigenetic clocks for the second cohort, specifically the rheumatoid arthritis (RA) dataset comprising 689 samples, with 335 (49%) being healthy control samples and 354 (51%) diagnosed with RA. A significant age difference between these two groups was identified by the researchers in this study (P value=0.0049), albeit with a small effect size of 1.2 years^58^. In contrast, Horvath’s study^12^ did not find a significant difference in negative age acceleration between diseased and healthy samples, as also evident in **Figure 7B** where we computed the DNAm age predictions and mean AgeAcc using Horvath’s algorithm. Interestingly, the average AgeAcc is slightly higher for the healthy cohort (Wilcoxon rank sum test P value = 0.0035). Conversely, for the other three algorithms, the diseased samples exhibit increased AgeAcc, with a marginal difference when using Hannum’s clock (0.3, Wilcoxon rank sum test P value = 0.0372), and a more pronounced difference in AgeAcc of 0.5 (Wilcoxon rank sum test P value < 0.0001) and 0.6 (Wilcoxon rank sum test P value = 0.0006) for PhenoAge and our methVAgE model, respectively. Notably, compared to PhenoAge, our model yields an average cohort AgeAcc > 1 for the RA samples, indicating that, on average, the RA subgroup is characterized by accelerated biological ageing.

**Figure 7:**
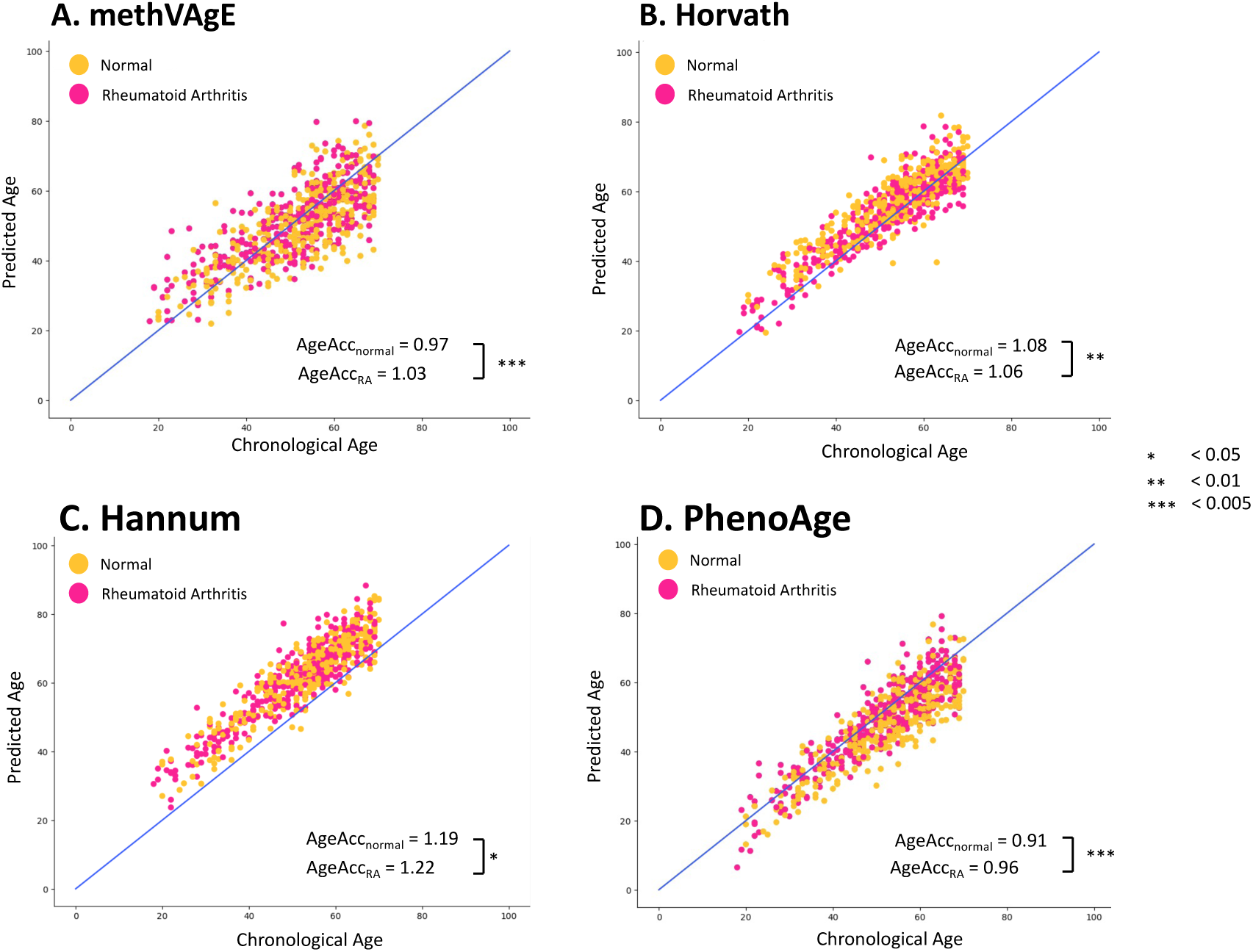
Model predictions of epigenetic clocks for the rheumatoid arthritis (RA) dataset. A. methVAgE, B. Horvath, C. Hannum and D. PhenoAge. Statistical significance between the AgeAcc of the diseased and normal samples was assessed using Wilcoxon rank sum test, with significance levels denoted by asterisks (*P value <0.05, **P value <0.01, and ***P value <0.005), indicating the level of significance in relation to the comparison of the two groups.

Moreover, in addition to evaluating AgeAcc, we investigated whether the latent space encoded by our VAE harbors biological signals that could serve as input for a classifier aimed at predicting age-related diseases. The rationale behind this approach is that the encoded feature space may exhibit differences between healthy and diseased samples, and a model could be trained to leverage these differences for disease classification. To test this hypothesis on the RA dataset, we trained a simple binary logistic regression model utilizing the VAE embeddings as input (as shown in **Figure 3**). The performance of this model on the training and test sets is presented in **Figure 8**. The left panel displays the ROC curves with the corresponding area under the curve (AUC) and F1 score, while the right panel depicts the confusion matrix. The results obtained with this simple model suggest that the VAE embeddings do indeed contain encoded biological signals that have the potential to discriminate between diseased and healthy samples.

**Figure 8:**
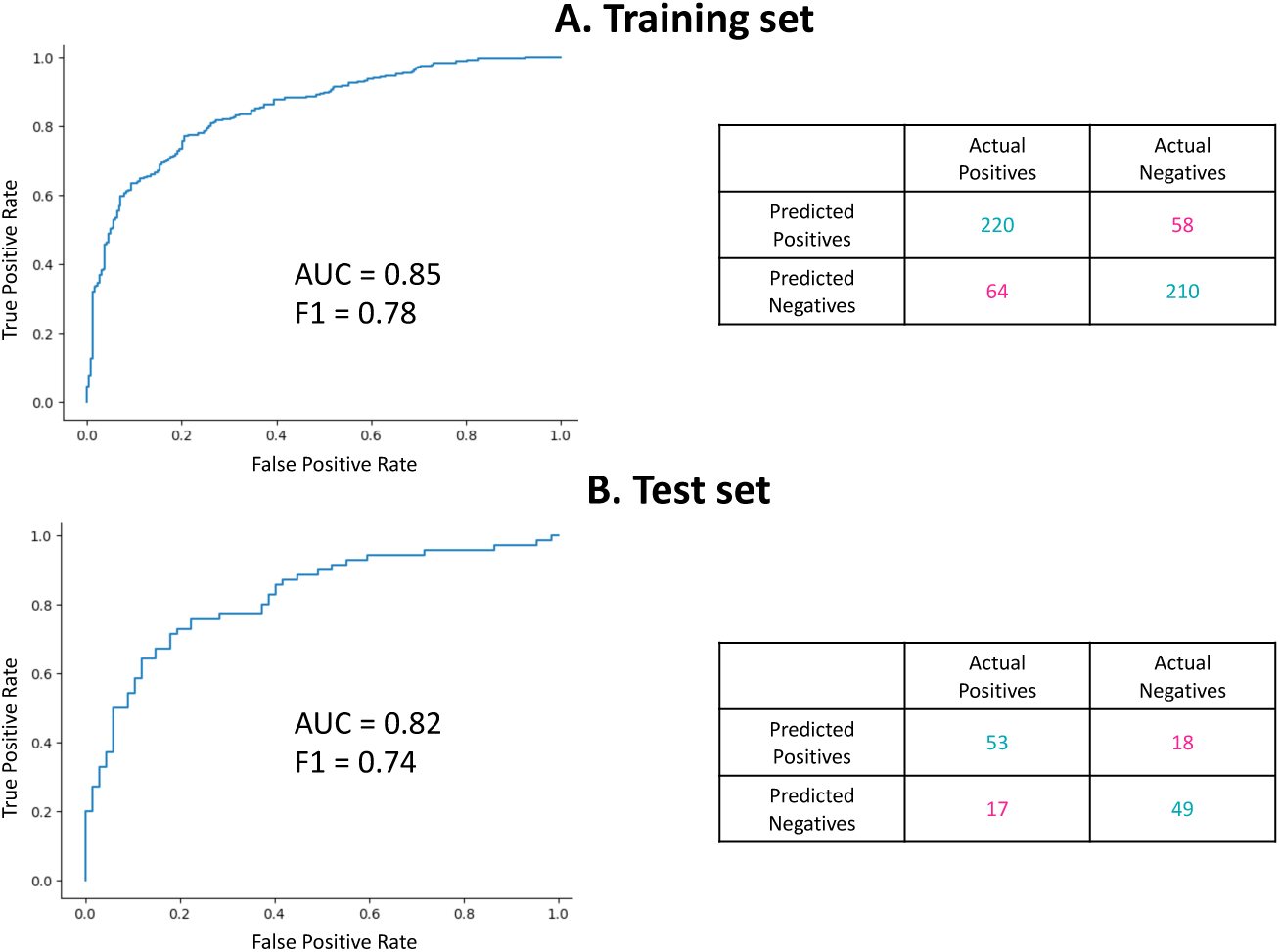
Performance of the binary logistic regression model using VAE embeddings as input for discriminating between diseased and healthy samples in the RA dataset. The left panel shows the Receiver Operating Characteristic (ROC) curves with the corresponding area under the curve (AUC) and F1 score, while the right panel displays the confusion matrix.

### Model interpretation

Based on the findings described above, we further conducted an unsupervised cluster analysis to determine if, when provided only with the VAE embeddings as input, such an analysis would independently reveal differences between the two groups. **Figure 9A** displays a 2D visualization of two different unsupervised cluster methods, namely t-SNE^61^ and UMAP^62^. Notably, most of the RA embeddings are clustered together in one group, while most normal samples are clustered in a distinct group. This observed independent clustering based solely on the VAE embeddings provides further evidence that the embeddings indeed contain relevant signals that may be associated with RA. In **Figure 9B** we present the same information, but with the samples colored according to their respective labels from the confusion matrix shown in **Figure 8**. It is noteworthy that the false positive samples are predominantly located within the RA cluster, while the false negative samples are mainly located outside the RA cluster. This observation suggests that the RA-related (biological) signals captured by the VAE embeddings are consistent with the classification results obtained from the logistic regression model.

**Figure 9:**
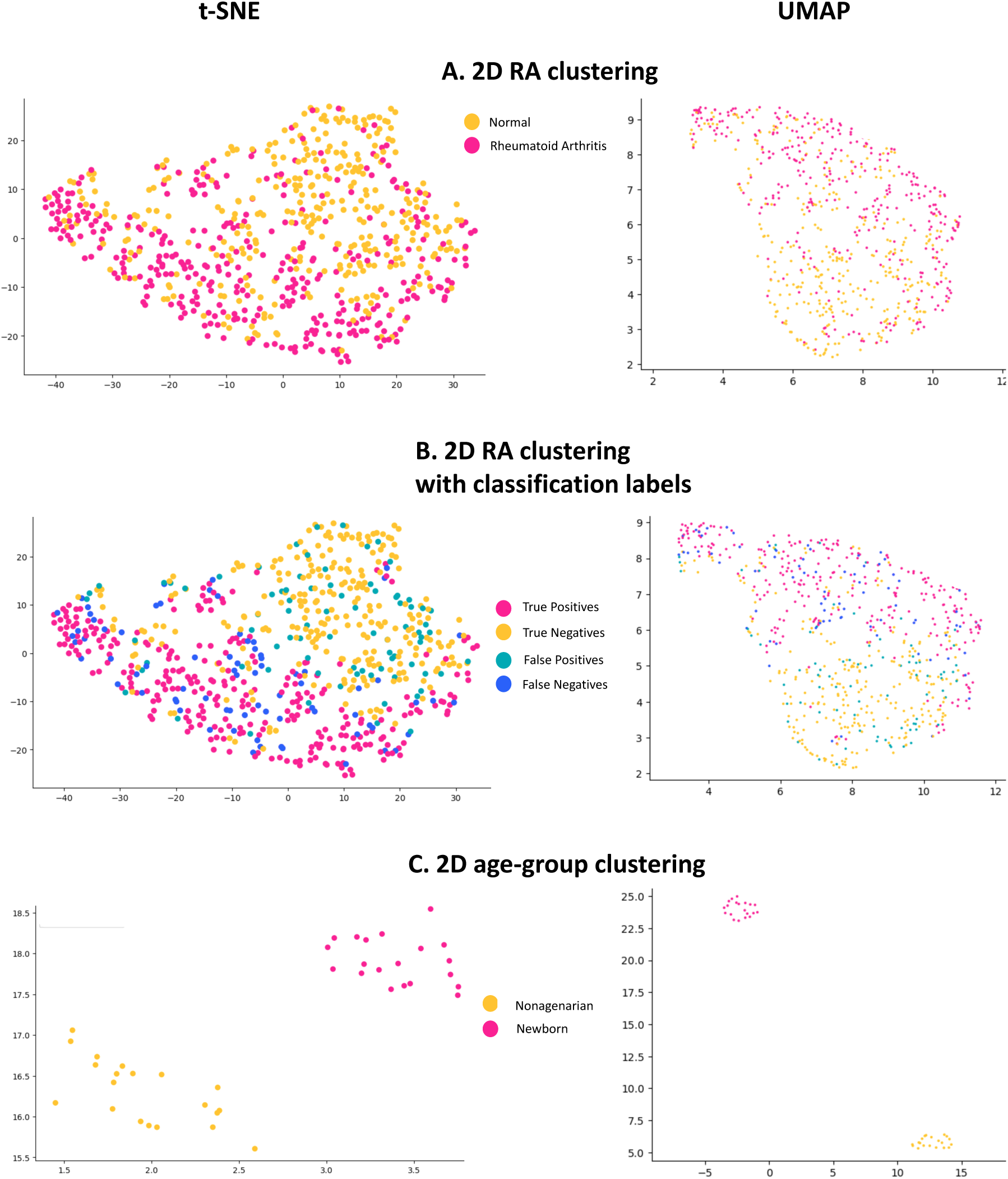
Unsupervised cluster analysis of VAE embeddings to reveal group differences. A. t-SNE and UMAP clustering results for RA dataset with the color indicating disease (RA vs normal). B. Same RA cluster analysis, but with the samples colored according to their labels from the confusion matrix shown in *Figure 8*. C. Cluster analysis for the GSE30870^46^ dataset comparing newborns versus nonagenarians.

Certainly, this type of analysis can be extended to explore differences among other groups of interest, such as different age-groups. In this regard, we performed a similar unsupervised clustering analysis using a dataset containing samples from both newborns and nonagenarians (GSE30870^46^, as described in **Table 1**), and the results are displayed in **Figure 9C**. Notably, the embeddings of these two age-groups are clustered in distinct groups, suggesting that the VAE embeddings contains relevant information that may be utilized to uncover mechanistic networks associated with the ageing process.

In addition to cluster analysis, the encoded feature representation in the VAE latent space can also be utilized to investigate biological mechanisms related to ageing and age-related disease. One approach to elucidate the biological signals is through pathway analysis of the model gradients, which allows for comparisons of pathway differences at both the individual and group levels. We applied this approach to compare RA and normal samples by backpropagating the VAE model and calculating gradients for each gene in the embedding. These individual gene gradients were then aggregated into Reactome pathways, and average pathway gradients were computed. Subsequently, these pathway gradients were compared between the two groups. **Supplementary Table 1** provides the full results, and **Table 4** highlights some of the top pathways that exhibited notable differences between the RA and healthy groups.

**Table 4:** Pathways that exhibited notable differences between the RA and healthy groups. The first column displays the pathway name, while the second column shows the ratio of pathway importance in the RA group compared to the normal group. The third column provides references on the involvement of each pathway in RA and/or ageing.

| Pathway | RA/normal ratio | References |
| --- | --- | --- |
| NEGATIVE FEEDBACK REGULATION OF MAPK PATHWAY | **10.91** | 64,65 |
| P75<sup>NTR</sup> NEGATIVELY REGULATES CELL CYCLE VIA SC1 | **2.00** | 68 |
| NF KB IS ACTIVATED AND SIGNALS SURVIVAL | **1.59** | 69 |
| N GLYCAN TRIMMING AND ELONGATION IN THE CIS GOLGI | **1.52** | 70 |
| FGFR3 LIGAND BINDING AND ACTIVATION | **1.50** | 71 |
| TNFR1 MEDIATED CERAMIDE PRODUCTION | **0.61** | 72 |
| SIGNALING BY MST1 | **0.53** | 73 |
| COMPETING ENDOGENOUS RNAS CERNAS REGULATE PTEN TRANSLATION | **0.53** | 74 |
| SYNTHESIS OF GDP MANNOSE | **0.005** | 75 |
| ACTIVATION OF THE AP1 FAMILY OF TRANSCRIPTION FACTORS | **<0.001** | 66,67 |

The first column displays the pathway name, while the second column shows the ratio of pathway importance in the RA group compared to the normal group. The third column provides references on the involvement of each pathway in RA. For example, the negative feedback regulation of the MAPK pathway showed a ratio of 10.91, indicating a nearly 11-fold higher presence in the RA group compared to the normal group. This finding is consistent with previous studies that have established the importance of the MAPK pathway in RA^64, 65^. On the other hand, the activation of the AP1 family of transcription factors was barely represented in the RA group but prominent in the normal group, which aligns with previous findings of diminished AP1 activity during ageing and in age-related diseases^66, 67^.

Again, a similar pathway analysis was performed to investigate potential differential representation of specific biological pathways in the comparison between newborns and nonagenarians. While **Supplementary Table 2** provides the full results, **Table 5** highlights some interesting pathways that exhibited notable differences between these two groups.

**Table 5:** Pathways that exhibited notable differences between the newborn and nonagenarian groups. The first column displays the pathway name, while the second column shows the ratio of pathway importance in the nonagenarian group compared to the newborn group. The third column provides references on the involvement of each pathway in ageing.

| Pathway | Nonagenarian / newborn ratio | References |
| --- | --- | --- |
| DISEASES ASSOCIATED WITH N GLYCOSYLATION OF PROTEINS | **201.82** | **76,77** |
| EICOSANOIDS | **6.69** | 80 |
| ELECTRIC TRANSMISSION ACROSS GAP JUNCTIONS | **4.97** | 81 |
| NR1H2 NR1H3 REGULATE GENE EXPRESSION LINKED TO GLUCONEOGENESIS | **4.03** | 79 |
| SYNTHESIS OF LEUKOTRIENES LT AND EOXINS EX | **2.23** | 82 |
| REGULATION OF BACH1 ACTIVITY | **2.22** | 83 |
| MECP2 REGULATES TRANSCRIPTION FACTORS | **0.37** | 84 |
| ERYTHROCYTES TAKE UP OXYGEN AND RELEASE CARBON DIOXIDE | **0.33** | 85 |
| CRISTAE FORMATION | **0.27** | 86 |
| POU5F1 OCT4 SOX2 NANOG ACTIVATE GENES RELATED TO PROLIFERATION | **0.19** | 78 |

For instance, the pathway “Diseases associated with N-glycosylation of proteins” is 200 times more important in nonagenarians compared to newborns. Ageing-associated glycosylation changes often resemble those observed in inflammatory conditions, and in fact, one of the most reproducible markers of both calendar and biological ageing is the presence of specific N-glycans^76, 77^. On the other hand, the involvement of transcription factors POU5F1, OCT4, SOX2, and NANOG is higher in newborns. Indeed, these transcription factors have been well-documented to be important in early differentiation and development^78^.

Lastly, there is an upregulated presence of gluconeogenesis in the nonagenarian population. Gluconeogenesis has been linked to healthy ageing, making this pathway especially relevant and not surprisingly linked to this nonagenarian population^79^.

## Discussion

Advances in DNAm arrays and data availability, has led to the creation of several methylation clocks, including those for humans but also other organisms. The best-known human DNAm clocks include those developed by Horvath^12^, Hannum^24^, Levine^13^, and Lu^25^, with the latter two being labeled as second-generation clocks due to their training on surrogate biomarkers reflecting ageing phenotypes to better predict biological age. Most methylation clocks are linear models of CpGs, selected through penalized or regularized regression methods like ElasticNet or Lasso. However, linear regression has limitations, such as high bias, inability to capture non-linear CpG interactions, and assumptions of additive contribution and constant methylation change rate over lifespan. DL has shown promise in DNAm age prediction with deep NN architectures outperforming linear regression models in chronological age prediction, being more resilient to noise, and capturing higher-order complex feature interactions^28, 33^. DL also allows for leveraging thousands of CpGs and interpreting their contribution using specific DL interpretation methods, making it a promising approach for epigenetic clock predictions, and understanding complex biological mechanisms in the context of ageing.

VAEs are a type of DL model that has shown great potential in various applications. VAEs are unsupervised generative models that use DL to learn data distributions without the need for accurate labels. They consist of encoding and decoding layers that compress the data into a lower-dimensional latent space and then reconstruct it back to its original dimension. The aim of the encoder phase is not only to reduce data dimensionality, but also to compress the data by removing redundant information while retaining important information. Unlike traditional autoencoders. VAEs can learn complex, lower-dimensional representations of the data due to the use of complex, non-linear activation functions. VAEs have already been successfully used in various biological applications.

In this study, we aimed to leverage the power of VAEs to construct a latent space in which methylation signals are encoded related to ageing that can subsequently be used to predict biological age. Using data from multiple cohorts (N = 2,716), two VAEs were trained, one with a KLD based on a Gaussian distribution and one with a distribution agnostic KLD implementation. The learned latent spaces of both implementations were subsequently used as input of an MLP to predict epigenetic age. Using MAE and r^2^ as performance metrics, the distribution-agnostic strategy achieved better epigenetic age predictions, with the final model having an MAE of 6.34 and r^2^ of 0.86 on the test set.

Models were next evaluated on two external datasets, i.e. one from an African American cohort (GENOA^57^, N=418) and a second dataset^58^ (N=689) containing both normal and RA samples. For the GENOA cohort, our methVAgeE framework consistently predicted DNAm ages that trended lower than the chronological ages at the cohort level (**Figure 6**). This observation is particularly intriguing in the context of previous findings that have demonstrated lower extrinsic DNAm ages in African Americans compared to whites. For Horvath and PhenoAge clocks this effect was less pronounced, suggesting that our framework may capture and emphasize these specific underlying biological signals associated with decelerated DNAm age. On the second cohort, i.e. the RA dataset, while Horvath’s clocks didn’t find accelerated ageing in the RA population compared to the healthy controls - in fact, AgeAcc was slightly higher for the healthy cohort (Wilcoxon rank sum test P value = 0.003) – Hannum’s, PhenoAge and methVAgE algorithms showed increased AgeAcc in diseased versus healthy samples, with marginal differences using Hannum’s clock (0.3 with P value = 0.037) and more pronounced differences of 0.5 (Wilcoxon rank sum test P value < 0.0001) and 0.6 (Wilcoxon rank sum test P value = 0.0006) for the PhenoAge and methVAgE models, respectively (**Figure 7**), confirming the original study’s results^58^. Compared to PhenoAge, the methVAgE model showed an average AgeAcc > 1 for the RA samples, thereby suggesting that the RA subgroup indeed experiences accelerated biological ageing.

In a next step, we investigated whether the latent space encoded by the VAE contains biological signals that could be utilized to predict age-related diseases. As a proof of concept, we trained a simple binary logistic regression model using the VAE embeddings as input, and disease labels as dependent variables. The results shown in **Figure 8** show that the VAE embeddings indeed contained encoded biological signals that have the potential to discriminate between RA and healthy samples (**Figure 8**). This is further evidenced by the subsequent cluster analysis (**Figure 9**). Most of the RA embeddings were found to be clustered together in one group, while most of the normal samples were clustered in another distinct group, consistent with the results obtained from the classification model. Taken together, this suggests that the RA embeddings contain meaningful signals that may be associated with RA. To unravel these biological signals, we performed a pathway analysis by backpropagating the model embeddings and fetching the CpG and corresponding gene gradients. After computing and comparing the average pathway gradients between the RA and healthy groups, valuable insights and potential associations with specific pathways underlying RA were found (**Table 4**). For example, the activation of the AP1 family of transcription factors was found to be barely represented in the RA group but prominent in the normal group, aligning with previous findings of diminished AP1 activity during ageing and in age-related diseases^66, 67^. The MAPK pathway on the other hand, was found to be highly important in the RA samples compared to normal samples, again consistent with previous studies emphasizing the importance of the MAPK pathway in RA^64, 65^. Overall, the pathway analysis of model gradients provides insights into the differential representation of specific biological pathways in the VAE latent space between the RA and normal groups, shedding light on potential pathways that may be involved in the pathogenesis of RA.

This type of analysis can be extended to explore differences other groups of interest, including other (age-related) diseases but also for healthy individuals such as different age-groups. As an example, we conducted the same type of analysis on a dataset containing data from both newborns and nonagenarians. Also here, the unsupervised clustering shows distinct groups (**Figure 9C**), suggesting that in addition to disease-networks, the VAE embeddings can potentially be utilized to uncover mechanistic networks associated with the ageing process itself. The subsequent pathway analysis indeed shows some notable differences between these two groups, with pathways upregulated in nonagenarians that have been already linked to ageing-associated processes, but also pathways more prominent in the newborn samples that have shown to be important for development (**Table 5**). This finding further underscores the potential utility of the VAE embeddings in capturing biologically meaningful signals in diverse contexts beyond disease datasets and highlights the versatility and applicability of this approach in exploring other populations of interest.

These results shows that VAE-based approaches may offer advantages in prediction biological age and understanding the complex dynamics of ageing. However, there are a couple of important remarks that come with our proposed methodology. Firstly, in the context of methylation data, the main strength of DL methods is their ability to simultaneously use information of thousands of CpGs, and to capture higher-order, complex CpG interactions. However, to reduce computational needs, we initially performed a prior dimensionality reduction by employing a bootstrapping ElasticNet procedure. This resulted in a reduced set of around 13k CpGs that were used as input features for training the VAE model. As such, the VAE does not leverage the full DNAm array during training. However, given the computational needs for training, such a prior subset procedure might be an advisable trade-off for computational resources versus efficiency. Here, as we were interested age-related health and diseae, we choose an ElasticNet selection with chronological age as dependent variable. And, although our strategy results in a significant larger subset of CpGs compared to other DNAm clocks (typically 3-2000 CpGs), the downside of this method is that it is linear by nature. Depending on the use case or research question, other methods and/or dependent variables can be chosen or can be even preferable. The same is true for our MLP model. Here we developed an MLP to predict DNAm age as a proof of concept for the utility of the VAE embeddings. But of course, other models or methods could also be chosen. While this model shows good performance for DNAm age and was mostly in line with PhenoAge (a second generation DNAm clock), the actual strength of this VAE framework is its applicable for multiple applications, exemplified by the RA classification and the pathway analysis. Indeed, an important remark that we want to make is that the potential is not limited to DNAm age. There has been a recent explosion in available clocks, and the debate which one is the best is – in our opinion – often subjective and mostly not relevant. In most cases it really drills down to the research purpose and being consistent. Often not the absolute values are important (and these can be highly biased by the used training set), but rather the relative differences between different samples or treatments/interventions or time points. The real added value of our framework lies when comparing between groups of interest and the potential to unravel biological meaningful insights. As a last remark, while we show the performance of our framework for a couple of different datasets, further investigations are warranted to fully assess the performance and utility of our proposed framework in different populations and settings.

In summary, the ability of DL methods to handle non-linear relationships and complex patterns in data holds great promise for interpreting various types of complex biological data within the context of ageing. As our understanding of ageing and its underlying mechanisms continues to advance, and as more complex and diverse data becomes available, such methods can play a critical role in extracting meaningful insights from these data. Here, we showed the potential of VAEs in combination with DNAm data to improve epigenetic age prediction and enhance our understanding of complex biological mechanisms underlying ageing and age-related diseases. Ultimately, the goal is to detect new biomarkers which can lead to the development of targeted interventions to delay or mitigate the effects of ageing and age-related diseases. However, further research and validation of these models in larger and diverse cohorts are needed to fully establish their utility and robustness in clinical and translational applications.

## Data Availability

All data used in the present study are publicly available and can be downloaded from the Gene Expression Omnibus (GEO) (GSE30870, GSE32149, GSE41169, GSE36064, GSE40279, GSE87571) and ArrayExpress (SATSA dataset - E-MTAB-7309). Details on the individual datasets and characteristics of the specific study cohort can be found in the original reference.

https://www.ncbi.nlm.nih.gov/geo/query/acc.cgi?acc=GSE30870

https://www.ncbi.nlm.nih.gov/geo/query/acc.cgi?acc=GSE32149

https://www.ncbi.nlm.nih.gov/geo/query/acc.cgi?acc=GSE41169

https://www.ncbi.nlm.nih.gov/geo/query/acc.cgi?acc=GSE36064

https://www.ncbi.nlm.nih.gov/geo/query/acc.cgi?acc=GSE40279

https://www.ncbi.nlm.nih.gov/geo/query/acc.cgi?acc=GSE87571

https://www.ebi.ac.uk/biostudies/arrayexpress/studies/E-MTAB-7309

## Notes

### Competing Interest Statement

The authors have declared no competing interest.

### Funding Statement

This study did not receive any funding.

